# Projected healthcare cost savings with transnasal vs. transoral upper gastrointestinal endoscopy in the United States

**DOI:** 10.1101/2023.11.27.23299075

**Authors:** Deepak Gupta, David Edelman, Amy Somerset, Shushovan Chakrabortty

**Author notes:** Corresponding Author: Dr Deepak Gupta Clinical Assistant Professor, Anesthesiology Wayne State University/Detroit Medical Center Box No 162, 3990 John R, Detroit, MI 48201, United States. Financial Interests: NONE; Conflicts of Interests: NONE. Anti-Plagiarism Software: Not Checked.

## Abstract

**Background:** Transoral esophagogastroduodenoscopy (EGD) requires sedation and monitored anesthesia care, while transnasal esophagoscopy (TNE) can be performed without sedation.

Despite this benefit, TNE is performed significantly less than EGD in the United States.

**Objectives:** The current project was designed to compare the differential prevalence of TNE and EGD with and without concurrent anesthesia billing among Medicare beneficiaries during 2018-2021.

**Materials and Methods:** The Public Use Files data for TNE, EGD, and anesthesia for EGD billed nationally among Medicare beneficiaries was exported from the Centers for Medicare & Medicaid Services (CMS) website for 2018-2021 and presented as cumulative data. Projections were made to quantify healthcare cost savings if the number of TNE services were at least 9% of the number of TNE+EGD services and at most 34% of the number of TNE+EGD services from 2018-2021.

**Results:** The key finding was that TNE was rarely performed and billed among Medicare beneficiaries when compared to EGD. In addition, roughly one-third of EGDs among Medicare beneficiaries might have been performed without concurrent anesthesia for EGD billing.

**Conclusion:** Secondary to the need for sedation and monitored anesthesia care, transoral upper gastrointestinal endoscopy is significantly more expensive than transnasal upper gastrointestinal endoscopy. Medicare could save millions of dollars annually if TNE replaced EGD.

## Introduction

Upper gastrointestinal endoscopy can be performed transorally or transnasally [1–10]. Transoral esophagogastroduodenoscopy (EGD) requires sedation and monitored anesthesia care, while transnasal esophagoscopy (TNE) can be performed without sedation. Despite this benefit, TNE is performed significantly less than EGD in the United States. In Japan, TNE is performed in up to 9% of cases in facility settings and up to 34% of cases in office settings [11]. The reasons for greater performance of TNE in eastern countries are unknown and may require comprehensive investigations or speculative discussions. In the United States, the Centers for Medicare & Medicaid Services (CMS) regularly reports Public Use Files (PUFs) about healthcare services rendered to its beneficiaries [12–14]. Therefore, the current project was designed to compare the differential prevalence of TNE and EGD with and without concurrent anesthesia billing among Medicare beneficiaries during 2018-2021.

## Materials and Methods

Institutional Review Board approval was not necessary as Public Use Files were used for data retrieval. The data for TNE, EGD, and anesthesia for EGD billed nationally among Medicare beneficiaries was exported from the CMS website for 2018-2021 (please see the links for the exports at Supplementary PDF). It was then presented as cumulative data in Tables. Projections were made to quantify healthcare cost savings if the number of TNE services were at least 9% of the number of TNE+EGD services and at most 34% of the number of TNE+EGD services from 2018-2021. These percentages were based on current percentages of TNE performed in Japanese facility and office settings, respectively.

## Results

The cumulative data by year for TNE, EGD, and anesthesia for EGD billed nationally are tabulated in the first six columns of Table 1. The subsequent eight columns include deduced national average amounts per provider (anesthesia provider or endoscopist) billing for their services and per service (anesthesia for EGD, EGD, or TNE) billed. Table 2 shows the deduced ratios of EGD by TNE in terms of number of providers, number of services and various types of amounts as well as deduced ratios of EGD with anesthesia by TNE based in terms of average amounts per service billed by the providers. As the number of services of anesthesia for EGD billed by anesthesia providers were significantly less than the number of services of EGD billed by endoscopists, it seemed appropriate to assume that the difference in these numbers likely represents EGDs performed with nonanesthesiologist/nurse-administered propofol/sedation and thus not billed by anesthesia providers. Therefore, Table 3 shows the reorganized data to represent those Medicare beneficiaries who might have been billed for EGD by endoscopists as well as anesthesia for EGD by anesthesia providers vs. those Medicare beneficiaries who might have been billed for EGD by endoscopists only. Current CMS data demonstrated that TNE was rare, with all EGD by TNE ratios being multi-thousands and thus representing incidence of all TNE data to be falling between 1 in 1000 to 1 in 10000 among all TNE+EGD data. Projections for each year were made by turning the incidences from rare (<0.1%) to incidence of 9% (common) and of 34% (very common) [15]; the corresponding savings for Medicare, in addition to the corresponding losses for endoscopists and anesthesia providers, were calculated based on the projected increased incidences of TNE (Tables 4-5).

**TABLE 1:** CUMULATIVE DATA ABOUT ANESTHESIA FOR ESOPHAGOGASTRODUODENOSCOPY, ESOPHAGOGASTRODUODENOSCOPY AND TRANSNASAL ESOPHAGOSCOPY BILLED DURING 2018-2021.

| Year | Number of Providers | Number of Services | Total Submitted Charge Amount | Total Medicare Allowed Amount | Total Medicare Payment Amount | Total Deductible Coinsurance Amount | Average Submitted Charge Amount Per Provider | Average Medicare Allowed Amount Per Provider | Average Medicare Payment Amount Per Provider | Average Deductible Coinsurance Amount Per Provider | Average Submitted Charge Amount Per Service | Average Medicare Allowed Amount Per Service | Average Medicare Payment Amount Per Service | Average Deductible Coinsurance Amount Per Service |
| --- | --- | --- | --- | --- | --- | --- | --- | --- | --- | --- | --- | --- | --- | --- |
| **ANESTHESIA FOR ESOPHAGOGASTRODUODENOSCOPY BILLED BY ANESTHESIA PROVIDERS NATIONALLY** |  |  |  |  |  |  |  |  |  |  |  |  |  |  |
| **2018** | 41,051 | 1,486,803 | 1,353,574,378 | 164,697,744 | 129,738,657 | 34,959,088 | 32,973 | 4,012 | 3,160 | 852 | 910 | 111 | 87 | 24 |
| **2019** | 42,512 | 1,510,975 | 1,429,327,702 | 169,007,556 | 133,041,609 | 35,965,947 | 33,622 | 3,976 | 3,130 | 846 | 946 | 112 | 88 | 24 |
| **2020** | 37,965 | 1,170,672 | 1,154,568,683 | 133,130,058 | 104,524,602 | 28,605,456 | 30,411 | 3,507 | 2,753 | 753 | 986 | 114 | 89 | 24 |
| **2021** | 40,188 | 1,247,273 | 1,289,592,402 | 138,876,944 | 109,495,607 | 29,381,337 | 32,089 | 3,456 | 2,725 | 731 | 1,034 | 111 | 88 | 24 |
| **ESOPHAGOGASTRODUODENOSCOPY BILLED BY ENDOSCOPISTS NATIONALLY** |  |  |  |  |  |  |  |  |  |  |  |  |  |  |
| **2018** | 28,192 | 2,265,300 | 2,528,475,096 | 367,146,113 | 286,740,615 | 80,405,498 | 89,688 | 13,023 | 10,171 | 2,852 | 1,116 | 162 | 127 | 35 |
| **2019** | 28,256 | 2,258,999 | 2,570,353,262 | 368,901,273 | 288,401,859 | 80,499,414 | 90,967 | 13,056 | 10,207 | 2,849 | 1,138 | 163 | 128 | 36 |
| **2020** | 25,504 | 1,775,647 | 2,057,269,789 | 288,220,499 | 224,858,249 | 63,362,249 | 80,665 | 11,301 | 8,817 | 2,484 | 1,159 | 162 | 127 | 36 |
| **2021** | 26,300 | 1,968,351 | 2,391,761,101 | 324,150,610 | 254,550,657 | 69,599,954 | 90,941 | 12,325 | 9,679 | 2,646 | 1,215 | 165 | 129 | 35 |
| **TRANSNASAL ESOPHAGOSCOPY BILLED BY ENDOSCOPISTS NATIONALLY** |  |  |  |  |  |  |  |  |  |  |  |  |  |  |
| **2018** | 24 | 898 | 657,642 | 138,109 | 104,665 | 33,444 | 27,402 | 5,755 | 4,361 | 1,394 | 732 | 154 | 117 | 37 |
| **2019** | 24 | 669 | 614,456 | 109,526 | 85,007 | 24,519 | 25,602 | 4,564 | 3,542 | 1,022 | 918 | 164 | 127 | 37 |
| **2020** | 20 | 546 | 415,731 | 93,704 | 72,060 | 21,644 | 20,787 | 4,685 | 3,603 | 1,082 | 761 | 172 | 132 | 40 |
| **2021** | 21 | 664 | 487,327 | 117,047 | 90,871 | 26,176 | 23,206 | 5,574 | 4,327 | 1,246 | 734 | 176 | 137 | 39 |

Source of Data: Centers for Medicare & Medicaid Services Public Use Files (See The Links To Raw Data In Supplementary PDF)

**TABLE 2:** COMPARATIVE RATIOS OF ESOPHAGOGASTRODUODENOSCOPY BY TRANSNASAL ESOPHAGOSCOPY AND ESOPHAGOGASTRODUODENOSCOPY WITH ANESTHESIA BY TRANSNASAL ESOPHAGOSCOPY DURING 2018-2021.

|  | Number of<br>Providers | Number of<br>Services | Total<br>Submitted<br>Charge<br>Amount | Total<br>Medicare<br>Allowed<br>Amount | Total<br>Medicare<br>Payment<br>Amount | Total<br>Deductible<br>Coinsurance<br>Amount | Average<br>Submitted<br>Charge<br>Amount Per<br>Service | Average<br>Medicare<br>Allowed<br>Amount Per<br>Service | Average<br>Medicare<br>Payment<br>Amount Per<br>Service | Average<br>Deductible<br>Coinsurance<br>Amount Per<br>Service | Average<br>Submitted<br>Charge<br>Amount Per<br>Service | Average<br>Medicare<br>Allowed<br>Amount Per<br>Service | Average<br>Medicare<br>Payment<br>Amount Per<br>Service | Average<br>Deductible<br>Coinsurance<br>Amount Per<br>Service |
| --- | --- | --- | --- | --- | --- | --- | --- | --- | --- | --- | --- | --- | --- | --- |
| Year | RATIO OF ESOPHAGOGASTRODUODENOSCOPY<br>BY TRANSNASAL ESOPHAGOSCOPY BILLED BY ENDOSCOPISTS NATIONALLY |  |  |  |  |  |  |  |  |  | RATIO OF ESOPHAGOGASTRODUODENOSCOPY WITH<br>ANESTHESIA BY TRANSNASAL ESOPHAGOSCOPY BILLED BY<br>ENDOSCOPISTS AND ANESTHESIA PROVIDERS NATIONALLY |  |  |  |
| 2018 | 1,175 | 2,523 | 3,845 | 2,658 | 2,740 | 2,404 | 2 | 1 | 1 | 1 | 3 | 2 | 2 | 2 |
| 2019 | 1,177 | 3,377 | 4,183 | 3,368 | 3,393 | 3,283 | 1 | 1 | 1 | 1 | 2 | 2 | 2 | 2 |
| 2020 | 1,275 | 3,252 | 4,949 | 3,076 | 3,120 | 2,928 | 2 | 1 | 1 | 1 | 3 | 2 | 2 | 2 |
| 2021 | 1,252 | 2,964 | 4,908 | 2,769 | 2,801 | 2,659 | 2 | 1 | 1 | 1 | 3 | 2 | 2 | 1 |

Source of Data: Centers for Medicare & Medicaid Services Public Use Files (See The Links To Raw Data In Supplementary PDF)

**TABLE 3:** REORGANIZED ESOPHAGOGASTRODUODENOSCOPY DATA INTO DATA ABOUT ESOPHAGOGASTRODUODENOSCOPY WITH ANESTHESIA AND ESOPHAGOGASTRODUODENOSCOPY WITHOUT ANESTHESIA BILLED DURING 2018-2021.

|  | Number of<br>Services | Total Submitted<br>Charge Amount | Total Medicare<br>Allowed Amount | Total Medicare<br>Payment Amount | Total Deductible<br>Coinsurance<br>Amount | Number of<br>Services | Total Submitted<br>Charge Amount | Total Medicare<br>Allowed Amount | Total Medicare<br>Payment Amount | Total Deductible<br>Coinsurance<br>Amount |
| --- | --- | --- | --- | --- | --- | --- | --- | --- | --- | --- |
| Year | ESOPHAGOGASTRODUODENOSCOPY WITH ANESTHESIA BILLED BY ENDOSCOPISTS AND ANESTHESIA<br>PROVIDERS NATIONALLY |  |  |  |  | ESOPHAGOGASTRODUODENOSCOPY WITHOUT ANESTHESIA BILLED BY ENDOSCOPISTS ONLY<br>NATIONALLY |  |  |  |  |
| 2018 | 1,486,803 | 3,013,109,255 | 405,669,776 | 317,937,486 | 87,732,290 | 778,497 | 868,940,218 | 126,174,082 | 98,541,786 | 27,632,295 |
| 2019 | 1,510,975 | 3,148,557,999 | 415,754,279 | 325,944,748 | 89,809,531 | 748,024 | 851,122,966 | 122,154,550 | 95,498,720 | 26,655,830 |
| 2020 | 1,170,672 | 2,510,912,673 | 323,151,874 | 252,772,118 | 70,379,756 | 604,975 | 700,925,798 | 98,198,683 | 76,610,734 | 21,587,949 |
| 2021 | 1,247,273 | 2,805,165,105 | 344,279,489 | 270,795,173 | 73,484,316 | 721,078 | 876,188,399 | 118,748,066 | 93,251,091 | 25,496,975 |

Source of Data: Centers for Medicare & Medicaid Services Public Use Files (See The Links To Raw Data In Supplementary PDF)

**TABLE 4:** PROJECTIONS IF AIMED TRANSNASAL ESOPHAGOSCOPY IS 9% (AND THUS AIMED ESOPHAGOGASTRODUODENOSCOPY IS 91%) OF CURRENT TOTAL ESOPHAGOGASTRODUODENOSCOPY PLUS TRANSNASAL ESOPHAGOSCOPY.

| Year | Projected Number of Services | Projected Total Submitted Charge Amount | Projected Total Medicare Allowed Amount | Projected Total Medicare Payment Amount | Projected Total Deductible Coinsurance Amount | Projected Savings In Total Submitted Charge Amount | Projected Savings In Total Medicare Allowed Amount | Projected Savings In Total Medicare Payment Amount | Projected Savings In Total Deductible Coinsurance Amount |
| --- | --- | --- | --- | --- | --- | --- | --- | --- | --- |
| PROJECTED TRANSNASAL ESOPHAGOSCOPY DATA |  |  |  |  |  | PROJECTED SAVINGS FOR MEDICARE |  |  |  |
| 2018 | 203,958 | 149,366,518 | 31,367,977 | 23,772,002 | 7,595,975 | 199,275,173 | 16,444,223 | 13,665,558 | 2,778,665 |
| 2019 | 203,370 | 186,789,372 | 33,294,914 | 25,841,259 | 7,453,655 | 172,718,475 | 15,081,443 | 12,060,082 | 3,021,360 |
| 2020 | 159,857 | 121,717,439 | 27,434,488 | 21,097,721 | 6,336,767 | 166,865,021 | 10,462,864 | 8,526,628 | 1,936,236 |
| 2021 | 177,211 | 130,060,155 | 31,238,110 | 24,252,182 | 6,985,928 | 200,618,894 | 10,409,278 | 8,491,099 | 1,918,179 |
| Year | Projected Number of Services | Projected Total Submitted Charge Amount | Projected Total Medicare Allowed Amount | Projected Total Medicare Payment Amount | Projected Total Deductible Coinsurance Amount | Projected Loss In Total Submitted Charge Amount | Projected Loss In Total Medicare Allowed Amount | Projected Loss In Total Medicare Payment Amount | Projected Loss In Total Deductible Coinsurance Amount |
| PROJECTED ESOPHAGOGASTRODUODENOSCOPY WITH ANESTHESIA DATA |  |  |  |  |  | PROJECTED LOSS FOR ANESTHESIA PROVIDERS NATIONALLY |  |  |  |
| 2018 | 1,353,527 | 2,743,016,365 | 369,305,836 | 289,437,804 | 79,868,032 | 121,333,408 | 14,763,384 | 11,629,677 | 3,133,707 |
| 2019 | 1,375,394 | 2,866,036,301 | 378,448,438 | 296,697,561 | 81,750,876 | 128,254,296 | 15,165,133 | 11,937,891 | 3,227,243 |
| 2020 | 1,065,639 | 2,285,633,134 | 294,158,630 | 230,093,358 | 64,065,272 | 103,588,111 | 11,944,453 | 9,377,966 | 2,566,487 |
| 2021 | 1,135,401 | 2,553,561,369 | 313,400,021 | 246,506,736 | 66,893,285 | 115,667,440 | 12,456,293 | 9,820,992 | 2,635,301 |
| Year | Projected Number of Services | Projected Total Submitted Charge Amount | Projected Total Medicare Allowed Amount | Projected Total Medicare Payment Amount | Projected Total Deductible Coinsurance Amount | Projected Loss In Total Submitted Charge Amount | Projected Loss In Total Medicare Allowed Amount | Projected Loss In Total Medicare Payment Amount | Projected Loss In Total Deductible Coinsurance Amount |
| PROJECTED ESOPHAGOGASTRODUODENOSCOPY WITHOUT ANESTHESIA DATA |  |  |  |  |  | PROJECTED LOSS FOR ENDOSCOPISTS NATIONALLY |  |  |  |
| 2018 | 708,713 | 791,049,058 | 114,863,930 | 89,708,573 | 25,155,357 | 77,941,765 | 1,680,839 | 2,035,881 | -355,042 |
| 2019 | 680,903 | 774,751,273 | 111,193,560 | 86,929,571 | 24,263,989 | 44,464,179 | -83,691 | 122,192 | -205,882 |
| 2020 | 550,697 | 638,038,609 | 89,388,279 | 69,737,205 | 19,651,074 | 63,276,910 | -1,481,589 | -851,338 | -630,251 |
| 2021 | 656,402 | 797,600,413 | 108,097,193 | 84,887,119 | 23,210,074 | 84,951,454 | -2,047,014 | -1,329,893 | -717,122 |

Projected From Sourced Data Per Centers for Medicare & Medicaid Services Public Use Files

**TABLE 5:** PROJECTIONS IF AIMED TRANSNASAL ESOPHAGOSCOPY IS 34% (AND THUS AIMED ESOPHAGOGASTRODUODENOSCOPY IS 66%) OF CURRENT TOTAL ESOPHAGOGASTRODUODENOSCOPY PLUS TRANSNASAL ESOPHAGOSCOPY.

| Year | Projected Number of Services | Projected Total Submitted Charge Amount | Projected Total Medicare Allowed Amount | Projected Total Medicare Payment Amount | Projected Total Deductible Coinsurance Amount | Projected Savings In Total Submitted Charge Amount | Projected Savings In Total Medicare Allowed Amount | Projected Savings In Total Medicare Payment Amount | Projected Savings In Total Deductible Coinsurance Amount |
| --- | --- | --- | --- | --- | --- | --- | --- | --- | --- |
| PROJECTED TRANSNASAL ESOPHAGOSCOPY DATA |  |  |  |  |  | PROJECTED SAVINGS FOR MEDICARE |  |  |  |
| 2018 | 770,507 | 564,273,511 | 118,501,246 | 89,805,340 | 28,695,906 | 755,265,275 | 62,324,626 | 51,793,313 | 10,531,314 |
| 2019 | 768,287 | 705,648,740 | 125,780,786 | 97,622,535 | 28,158,251 | 654,075,474 | 57,112,603 | 45,670,876 | 11,441,727 |
| 2020 | 603,906 | 459,821,437 | 103,641,398 | 79,702,501 | 23,938,896 | 631,967,546 | 39,625,984 | 32,292,881 | 7,333,103 |
| 2021 | 669,465 | 491,338,363 | 118,010,636 | 91,619,354 | 26,391,282 | 759,989,527 | 39,432,690 | 32,166,195 | 7,266,495 |
| Year | Projected Number of Services | Projected Total Submitted Charge Amount | Projected Total Medicare Allowed Amount | Projected Total Medicare Payment Amount | Projected Total Deductible Coinsurance Amount | Projected Loss In Total Submitted Charge Amount | Projected Loss In Total Medicare Allowed Amount | Projected Loss In Total Medicare Payment Amount | Projected Loss In Total Deductible Coinsurance Amount |
| PROJECTED ESOPHAGOGASTRODUODENOSCOPY WITH ANESTHESIA DATA |  |  |  |  |  | PROJECTED LOSS FOR ANESTHESIA PROVIDERS NATIONALLY |  |  |  |
| 2018 | 981,679 | 1,989,440,441 | 267,848,189 | 209,921,924 | 57,926,265 | 459,861,147 | 55,954,142 | 44,077,199 | 11,876,943 |
| 2019 | 997,539 | 2,078,663,691 | 274,479,087 | 215,187,242 | 59,291,844 | 485,692,045 | 57,429,535 | 45,208,143 | 12,221,392 |
| 2020 | 772,881 | 1,657,711,944 | 213,345,819 | 166,880,897 | 46,464,922 | 392,319,037 | 45,237,202 | 35,517,152 | 9,720,050 |
| 2021 | 823,478 | 1,852,033,520 | 227,301,114 | 178,785,105 | 48,516,009 | 438,174,298 | 47,187,241 | 37,204,128 | 9,983,113 |
| Year | Projected Number of Services | Projected Total Submitted Charge Amount | Projected Total Medicare Allowed Amount | Projected Total Medicare Payment Amount | Projected Total Deductible Coinsurance Amount | Projected Loss In Total Submitted Charge Amount | Projected Loss In Total Medicare Allowed Amount | Projected Loss In Total Medicare Payment Amount | Projected Loss In Total Deductible Coinsurance Amount |
| PROJECTED ESOPHAGOGASTRODUODENOSCOPY WITHOUT ANESTHESIA DATA |  |  |  |  |  | PROJECTED LOSS FOR ENDOSCOPISTS NATIONALLY |  |  |  |
| 2018 | 514,012 | 573,727,889 | 83,307,905 | 65,063,361 | 18,244,545 | 295,404,127 | 6,370,484 | 7,716,114 | -1,345,630 |
| 2019 | 493,842 | 561,907,516 | 80,645,879 | 63,047,821 | 17,598,058 | 168,383,429 | -316,932 | 462,733 | -779,665 |
| 2020 | 399,406 | 462,753,277 | 64,831,060 | 50,578,632 | 14,252,427 | 239,648,508 | -5,611,218 | -3,224,270 | -2,386,947 |
| 2021 | 476,072 | 578,479,421 | 78,400,162 | 61,566,482 | 16,833,680 | 321,815,229 | -7,754,551 | -5,037,933 | -2,716,618 |

Projected From Sourced Data Per Centers for Medicare & Medicaid Services Public Use Files

## Discussion

The key finding of the current project based on CMS PUFs data for the 2018-2021 period was that TNE was rarely performed and billed among Medicare beneficiaries when compared to EGD (Table 2). In addition, roughly one-third of EGDs among Medicare beneficiaries might have been performed without concurrent anesthesia for EGD billing (Table 3). There is potential for these procedures to have been performed with nonanesthesiologist/nurse-administered propofol/sedation [16–20]. The cost of monitored anesthesia care is significant, and TNE is safely completed in its absence. Medicare could save millions of dollars annually if TNE replaced EGD (Tables 4-5). If TNE became common, as it is in Japanese facility-based cases, a 9% revenue loss would be seen for anesthesia providers, compared to a 34% revenue loss if TNE were very common, as it is in Japanese office-based cases (please see additional projections in Supplementary Excel File). Paradoxically, endoscopists might experience increased revenue as reflected by their negative losses in Tables 4-5 which may be due to the fact that average payment amount per service may be equivalent whether endoscopists perform EGD or TNE (Table 1) thus meaning projected Medicare savings being disproportionately due to the losses to anesthesia providers for not getting to perform and bill anesthesia for EGD. Moreover, as compared to anesthesia for EGD having a unique billing code, both EGD and TNE have dual codes depending on whether or not a biopsy is performed. Therefore, the projected revenue increase (negative losses) for individual endoscopists may have gotten undervalued in Tables 4-5 considering that data of providers performing EGD or TNE with as well as without biopsy might have been counted twice. This duplicity in counting endoscopists could not be addressed during our current analysis (please see EGD or TNE links for their exports at Supplementary PDF).

With significant cost saving projections, CMS, and commercial third-party payers thereafter, may expect the adoption of TNE by endoscopists, similar to countries such as Japan, where TNE is most commonly performed. Patients, endoscopists and anesthesia providers may have multiple concerns with this shift in practice. There is a question of patient comfort with TNE compared to EGD, which could result in poor patient satisfaction and incomplete procedures. Given that TNE is rarely performed in the United States, proceduralists will require additional training that is not routinely provided during current residencies and fellowships. Certainly, adequate training in TNE must be achieved prior to ubiquitous adoption of this procedure, as healthcare costs would actually increase if subsequent EGD were required for inadequate TNE.

Despite these obstacles, there are benefits beyond cost which may be seen with performing TNE. There are risks associated with the administration of sedation or anesthesia, which may include the patient’s inability to maintain and protect the airway, adequate spontaneous ventilation, and stable cardiovascular function [21–22]. High-risk patients, such as those with obesity or cardiopulmonary diseases, can undergo endoscopy in a safer manner when eliminating the need for anesthesia. Complications related to sedation or anesthesia itself increase the intraoperative duration of procedures and increase overall risk to the patient. Omission of monitored anesthesia care will obviate the need for additional time required for anesthesia preparation, induction, and recovery. Post-procedural monitoring in a Post-Anesthesia Care Unit (PACU) would be unnecessary, which reduces the need for trained staff and resources and further drives down costs. In addition, the required long-duration pre-procedural fasting instructions would no longer be applicable in the absence of sedation, and satisfaction would be greater with minimal disruption of typical patient diet routines [23–25]. Lastly, patients would not require an escort for transportation as they can safely transport themselves to and from the Endoscopy Center [26]. The associated benefits of TNE could actually result in additional procedures and increased revenue. Patient satisfaction may lead to a greater referral base and an increased number of cases. With the reduction of pre-, intra-, and post-procedural time associated with sedation and anesthesia, procedures can be performed more rapidly with decreased turnover time. This can translate into a greater number of endoscopies being completed.

## Conclusion

Secondary to the need for sedation and monitored anesthesia care, transoral upper gastrointestinal endoscopy is significantly more expensive than transnasal upper gastrointestinal endoscopy. TNE could save millions for CMS if endoscopists adopted this form of endoscopy and altered their practice of nearly-exclusive transoral EGD. These savings would be primarily due to the absence of costs related to sedation or anesthesia. While anesthesia providers would see a significant reduction in billable income, endoscopists may actually increase their revenues, as current payments for TNE per service are almost equivalent to current payments for EGD per service. Eventually, endoscopists may be able to convince public and private insurers to improve payments for TNE given the significant savings with elimination of sedation and monitored anesthesia care.

## Supporting information

Non-Human Participant Research

## Data Availability

All data produced in the present work are contained in the manuscript and its Supplementary Excel File
All raw data used are available online at CMS website links as detailed in Supplementary PDF of the manuscript

## Acknowledgment

The authors are indebted to the Centers for Medicare & Medicaid Services and their website for open access public domain Public Use Files data.

