## Supplementary PDF for "Projected healthcare cost savings with transnasal vs. transoral upper gastrointestinal endoscopy in the United States"

2018 ANESTHESIA FOR ESOPHAGOGASTRODUODENOSCOPY RAW DATA LINK

https://data.cms.gov/provider-summary-by-type-of-service/medicare-physician-other-practitioners/medicare-physician-other-practitioners-by-provider-and-service/data/2018?query=%7B%22filters%22%3A%7B%22list%22%3A%5B%7B%22conjunction%22%3A%7B%22value%22%3A%22OR%22%7D%2C%22conditions%22%3A%5B%7B%22column%22%3A%7B%22value%22%3A%22HCPCS_Cd%22%7D%2C%22comparator%22%3A%7B%22value%22%3A%22%3D%22%7D%2C%22filterValue%22%3A%5B%2200731%22%5D%7D%5D%7D%5D%2C%22rootConjunction%22%3A%7B%22value%22%3A%22AND%22%7D%7D%2C%22keywords%22%3A%22%22%2C%22offset%22%3A0%2C%22limit%22%3A10%2C%22sort%22%3A%7B%22sortBy%22%3Anull%2C%22sortOrder%22%3Anull%7D%2C%22columns%22%3A%5B%22HCPCS_Desc%22%2C%22Tot_Srvcs%22%2C%22Avg_Sbmtd_Chrg%22%2C%22Avg_Mdcr_Alowd_Amt%22%2C%22Avg_Mdcr_Pymt_Amt%22%5D%7D

2019 ANESTHESIA FOR ESOPHAGOGASTRODUODENOSCOPY RAW DATA LINK

https://data.cms.gov/provider-summary-by-type-of-service/medicare-physician-other-practitioners/medicare-physician-other-practitioners-by-provider-and-service/data/2019?query=%7B%22filters%22%3A%7B%22list%22%3A%5B%7B%22conjunction%22%3A%7B%22value%22%3A%22OR%22%7D%2C%22conditions%22%3A%5B%7B%22column%22%3A%7B%22value%22%3A%22HCPCS_Cd%22%7D%2C%22comparator%22%3A%7B%22value%22%3A%22%3D%22%7D%2C%22filterValue%22%3A%5B%2200731%22%5D%7D%5D%7D%5D%2C%22rootConjunction%22%3A%7B%22value%22%3A%22AND%22%7D%7D%2C%22keywords%22%3A%22%22%2C%22offset%22%3A0%2C%22limit%22%3A10%2C%22sort%22%3A%7B%22sortBy%22%3Anull%2C%22sortOrder%22%3Anull%7D%2C%22columns%22%3A%5B%22HCPCS_Desc%22%2C%22Tot_Srvcs%22%2C%22Avg_Sbmtd_Chrg%22%2C%22Avg_Mdcr_Alowd_Amt%22%2C%22Avg_Mdcr_Pymt_Amt%22%5D%7D

2020 ANESTHESIA FOR ESOPHAGOGASTRODUODENOSCOPY RAW DATA LINK

https://data.cms.gov/provider-summary-by-type-of-service/medicare-physician-other-practitioners/medicare-physician-other-practitioners-by-provider-and-service/data/2020?query=%7B%22filters%22%3A%7B%22list%22%3A%5B%7B%22conjunction%22%3A%7B%22value%22%3A%22OR%22%7D%2C%22conditions%22%3A%5B%7B%22column%22%3A%7B%22value%22%3A%22HCPCS_Cd%22%7D%2C%22comparator%22%3A%7B%22value%22%3A%22%3D%22%7D%2C%22filterValue%22%3A%5B%2200731%22%5D%7D%5D%7D%5D%2C%22rootConjunction%22%3A%7B%22value%22%3A%22AND%22%7D%7D%2C%22keywords%22%3A%22%22%2C%22offset%22%3A0%2C%22limit%22%3A10%2C%22sort%22%3A%7B%22sortBy%22%3Anull%2C%22sortOrder%22%3Anull%7D%2C%22columns%22%3A%5B%22HCPCS_Desc%22%2C%22Tot_Srvcs%22%2C%22Avg_Sbmtd_Chrg%22%2C%22Avg_Mdcr_Alowd_Amt%22%2C%22Avg_Mdcr_Pymt_Amt%22%5D%7D

2021 ANESTHESIA FOR ESOPHAGOGASTRODUODENOSCOPY RAW DATA LINK

https://data.cms.gov/provider-summary-by-type-of-service/medicare-physician-other-practitioners/medicare-physician-other-practitioners-by-provider-and-service/data/?query=%7B%22filters%22%3A%7B%22list%22%3A%5B%7B%22conjunction%22%3A%7B%22value%22%3A%22OR%22%7D%2C%22conditions%22%3A%5B%7B%22column%22%3A%7B%22value%22%3A%22HCPCS_Cd%22%7D%2C%22comparator%22%3A%7B%22value%22%3A%22%3D%22%7D%2C%22filterValue%22%3A%5B%2200731%22%5D%7D%5D%7D%5D%2C%22rootConjunction%22%3A%7B%22value%22%3A%22AND%22%7D%7D%2C%22keywords%22%3A%22%22%2C%22offset%22%3A0%2C%22limit%22%3A10%2C%22sort%22%3A%7B%22sortBy%22%3Anull%2C%22sortOrder%22%3Anull%7D%2C%22columns%22%3A%5B%22HCPCS_Desc%22%2C%22Tot_Srvcs%22%2C%22Avg_Sbmtd_Chrg%22%2C%22Avg_Mdcr_Alowd_Amt%22%2C%22Avg_Mdcr_Pymt_Amt%22%5D%7D

2018 ESOPHAGOGASTRODUODENOSCOPY RAW DATA LINK

https://data.cms.gov/provider-summary-by-type-of-service/medicare-physician-other-
practitioners/medicare-physician-other-practitioners-by-provider-and-
service/data/2018?query=%7B%22filters%22%3A%7B%22list%22%3A%5B%7B%22conjunction%22%3A
%7B%22value%22%3A%22OR%22%7D%2C%22conditions%22%3A%5B%7B%22column%22%3A%7B%22
value%22%3A%22HCPCS_Cd%22%7D%2C%22comparator%22%3A%7B%22value%22%3A%22%3D%22%
7D%2C%22filterValue%22%3A%5B%2243235%22%5D%7D%2C%7B%22column%22%3A%7B%22value%2
2%3A%22HCPCS_Cd%22%7D%2C%22comparator%22%3A%7B%22value%22%3A%22%3D%22%7D%2C%
22filterValue%22%3A%5B%2243239%22%5D%7D%5D%7D%5D%2C%22rootConjunction%22%3A%7B%2
2value%22%3A%22AND%22%7D%7D%2C%22keywords%22%3A%22%22%2C%22offset%22%3A0%2C%2
2limit%22%3A10%2C%22sort%22%3A%7B%22sortBy%22%3Anull%2C%22sortOrder%22%3Anull%7D%2
C%22columns%22%3A%5B%22HCPCS_Desc%22%2C%22Tot_Srvcs%22%2C%22Avg_Sbmtd_Chrg%22%2C
%22Avg_Mdcr_Alowd_Amt%22%2C%22Avg_Mdcr_Pymt_Amt%22%5D%7D

2019 ESOPHAGOGASTRODUODENOSCOPY RAW DATA LINK

https://data.cms.gov/provider-summary-by-type-of-service/medicare-physician-other-practitioners/medicare-physician-other-practitioners-by-provider-and-service/data/2019?query=%7B%22filters%22%3A%7B%22list%22%3A%5B%7B%22conjunction%22%3A%7B%22value%22%3A%22OR%22%7D%2C%22conditions%22%3A%5B%7B%22column%22%3A%7B%22value%22%3A%22HCPCS_Cd%22%7D%2C%22comparator%22%3A%7B%22value%22%3A%22%3D%22%7D%2C%22filterValue%22%3A%5B%2243235%22%5D%7D%2C%7B%22column%22%3A%7B%22value%22%3A%22HCPCS_Cd%22%7D%2C%22comparator%22%3A%7B%22value%22%3A%22%3D%22%7D%2C%22filterValue%22%3A%5B%2243239%22%5D%7D%5D%7D%5D%2C%22rootConjunction%22%3A%7B%22value%22%3A%22AND%22%7D%7D%2C%22keywords%22%3A%22%22%2C%22offset%22%3A0%2C%22limit%22%3A10%2C%22sort%22%3A%7B%22sortBy%22%3Anull%2C%22sortOrder%22%3Anull%7D%2C%22columns%22%3A%5B%22HCPCS_Desc%22%2C%22Tot_Srvcs%22%2C%22Avg_Sbmtd_Chrg%22%2C%22Avg_Mdcr_Alowd_Amt%22%2C%22Avg_Mdcr_Pymt_Amt%22%5D%7D

2020 ESOPHAGOGASTRODUODENOSCOPY RAW DATA LINK

https://data.cms.gov/provider-summary-by-type-of-service/medicare-physician-other-practitioners/medicare-physician-other-practitioners-by-provider-and-service/data/2020?query=%7B%22filters%22%3A%7B%22list%22%3A%5B%7B%22conjunction%22%3A%7B%22value%22%3A%22OR%22%7D%2C%22conditions%22%3A%5B%7B%22column%22%3A%7B%22value%22%3A%22HCPCS_Cd%22%7D%2C%22comparator%22%3A%7B%22value%22%3A%22%3D%22%7D%2C%22filterValue%22%3A%5B%2243235%22%5D%7D%2C%7B%22column%22%3A%7B%22value%22%3A%22HCPCS_Cd%22%7D%2C%22comparator%22%3A%7B%22value%22%3A%22%3D%22%7D%2C%22filterValue%22%3A%5B%2243239%22%5D%7D%5D%7D%5D%2C%22rootConjunction%22%3A%7B%22value%22%3A%22AND%22%7D%7D%2C%22keywords%22%3A%22%22%2C%22offset%22%3A0%2C%22limit%22%3A10%2C%22sort%22%3A%7B%22sortBy%22%3Anull%2C%22sortOrder%22%3Anull%7D%2C%22columns%22%3A%5B%22HCPCS_Desc%22%2C%22Tot_Srvcs%22%2C%22Avg_Sbmtd_Chrg%22%2C%22Avg_Mdcr_Alowd_Amt%22%2C%22Avg_Mdcr_Pymt_Amt%22%5D%7D

2021 ESOPHAGOGASTRODUODENOSCOPY RAW DATA LINK

https://data.cms.gov/provider-summary-by-type-of-service/medicare-physician-other-practitioners/medicare-physician-other-practitioners-by-provider-and-service/data/?query=%7B%22filters%22%3A%7B%22list%22%3A%5B%7B%22conjunction%22%3A%7B%22value%22%3A%22OR%22%7D%2C%22conditions%22%3A%5B%7B%22column%22%3A%7B%22value%22%3A%22HCPCS_Cd%22%7D%2C%22comparator%22%3A%7B%22value%22%3A%22%3D%22%7D%2C%22filterValue%22%3A%5B%2243235%22%5D%7D%2C%7B%22column%22%3A%7B%22value%22%3A%22HCPCS_Cd%22%7D%2C%22comparator%22%3A%7B%22value%22%3A%22%3D%22%7D%2C%22filterValue%22%3A%5B%2243239%22%5D%7D%5D%7D%5D%2C%22rootConjunction%22%3A%7B%22value%22%3A%22AND%22%7D%7D%2C%22keywords%22%3A%22%22%2C%22offset%22%3A0%2C%22limit%22%3A10%2C%22sort%22%3A%7B%22sortBy%22%3Anull%2C%22sortOrder%22%3Anull%7D%2C%22columns%22%3A%5B%22HCPCS_Desc%22%2C%22Tot_Srvcs%22%2C%22Avg_Sbmtd_Chrg%22%2C%22Avg_Mdcr_Alowd_Amt%22%2C%22Avg_Mdcr_Pymt_Amt%22%5D%7D

2018 TRANSNASAL ESOPHAGOSCOPY RAW DATA LINK

https://data.cms.gov/provider-summary-by-type-of-service/medicare-physician-other-practitioners/medicare-physician-other-practitioners-by-provider-and-service/data/2018?query=%7B%22filters%22%3A%7B%22list%22%3A%5B%7B%22conjunction%22%3A%7B%22value%22%3A%22OR%22%7D%2C%22conditions%22%3A%5B%7B%22column%22%3A%7B%22value%22%3A%22HCPCS_Cd%22%7D%2C%22comparator%22%3A%7B%22value%22%3A%22%3D%22%7D%2C%22filterValue%22%3A%5B%2243197%22%5D%7D%2C%7B%22column%22%3A%7B%22value%22%3A%22HCPCS_Cd%22%7D%2C%22comparator%22%3A%7B%22value%22%3A%22%3D%22%7D%2C%22filterValue%22%3A%5B%2243198%22%5D%7D%5D%7D%5D%2C%22rootConjunction%22%3A%7B%22value%22%3A%22AND%22%7D%7D%2C%22keywords%22%3A%22%22%2C%22offset%22%3A0%2C%22limit%22%3A10%2C%22sort%22%3A%7B%22sortBy%22%3Anull%2C%22sortOrder%22%3Anull%7D%2C%22columns%22%3A%5B%22HCPCS_Desc%22%2C%22Tot_Srvcs%22%2C%22Avg_Sbmtd_Chrg%22%2C%22Avg_Mdcr_Alowd_Amt%22%2C%22Avg_Mdcr_Pymt_Amt%22%5D%7D

2019 TRANSNASAL ESOPHAGOSCOPY RAW DATA LINK

https://data.cms.gov/provider-summary-by-type-of-service/medicare-physician-other-practitioners/medicare-physician-other-practitioners-by-provider-and-service/data/2019?query=%7B%22filters%22%3A%7B%22list%22%3A%5B%7B%22conjunction%22%3A%7B%22value%22%3A%22OR%22%7D%2C%22conditions%22%3A%5B%7B%22column%22%3A%7B%22value%22%3A%22HCPCS_Cd%22%7D%2C%22comparator%22%3A%7B%22value%22%3A%22%3D%22%7D%2C%22filterValue%22%3A%5B%2243197%22%5D%7D%2C%7B%22column%22%3A%7B%22value%22%3A%22HCPCS_Cd%22%7D%2C%22comparator%22%3A%7B%22value%22%3A%22%3D%22%7D%2C%22filterValue%22%3A%5B%2243198%22%5D%7D%5D%7D%5D%2C%22rootConjunction%22%3A%7B%22value%22%3A%22AND%22%7D%7D%2C%22keywords%22%3A%22%22%2C%22offset%22%3A0%2C%22limit%22%3A10%2C%22sort%22%3A%7B%22sortBy%22%3Anull%2C%22sortOrder%22%3Anull%7D%2C%22columns%22%3A%5B%22HCPCS_Desc%22%2C%22Tot_Srvcs%22%2C%22Avg_Sbmtd_Chrg%22%2C%22Avg_Mdcr_Alowd_Amt%22%2C%22Avg_Mdcr_Pymt_Amt%22%5D%7D

2020 TRANSNASAL ESOPHAGOSCOPY RAW DATA LINK

https://data.cms.gov/provider-summary-by-type-of-service/medicare-physician-other-practitioners/medicare-physician-other-practitioners-by-provider-and-service/data/2020?query=%7B%22filters%22%3A%7B%22list%22%3A%5B%7B%22conjunction%22%3A%7B%22value%22%3A%22OR%22%7D%2C%22conditions%22%3A%5B%7B%22column%22%3A%7B%22value%22%3A%22HCPCS_Cd%22%7D%2C%22comparator%22%3A%7B%22value%22%3A%22%3D%22%7D%2C%22filterValue%22%3A%5B%2243197%22%5D%7D%2C%7B%22column%22%3A%7B%22value%22%3A%22HCPCS_Cd%22%7D%2C%22comparator%22%3A%7B%22value%22%3A%22%3D%22%7D%2C%22filterValue%22%3A%5B%2243198%22%5D%7D%5D%7D%5D%2C%22rootConjunction%22%3A%7B%22value%22%3A%22AND%22%7D%7D%2C%22keywords%22%3A%22%22%2C%22offset%22%3A0%2C%22limit%22%3A10%2C%22sort%22%3A%7B%22sortBy%22%3Anull%2C%22sortOrder%22%3Anull%7D%2C%22columns%22%3A%5B%22HCPCS_Desc%22%2C%22Tot_Srvcs%22%2C%22Avg_Sbmtd_Chrg%22%2C%22Avg_Mdcr_Alowd_Amt%22%2C%22Avg_Mdcr_Pymt_Amt%22%5D%7D

2021 TRANSNASAL ESOPHAGOSCOPY RAW DATA LINK

https://data.cms.gov/provider-summary-by-type-of-service/medicare-physician-other-practitioners/medicare-physician-other-practitioners-by-provider-and-service/data/?query=%7B%22filters%22%3A%7B%22list%22%3A%5B%7B%22conjunction%22%3A%7B%22value%22%3A%22OR%22%7D%2C%22conditions%22%3A%5B%7B%22column%22%3A%7B%22value%22%3A%22HCPCS_Cd%22%7D%2C%22comparator%22%3A%7B%22value%22%3A%22%3D%22%7D%2C%22filterValue%22%3A%5B%2243197%22%5D%7D%2C%7B%22column%22%3A%7B%22value%22%3A%22HCPCS_Cd%22%7D%2C%22comparator%22%3A%7B%22value%22%3A%22%3D%22%7D%2C%22filterValue%22%3A%5B%2243198%22%5D%7D%5D%7D%5D%2C%22rootConjunction%22%3A%7B%22value%22%3A%22AND%22%7D%7D%2C%22keywords%22%3A%22%22%2C%22offset%22%3A0%2C%22limit%22%3A10%2C%22sort%22%3A%7B%22sortBy%22%3Anull%2C%22sortOrder%22%3Anull%7D%2C%22columns%22%3A%5B%22HCPCS_Desc%22%2C%22Tot_Srvcs%22%2C%22Avg_Sbmtd_Chrg%22%2C%22Avg_Mdcr_Alowd_Amt%22%2C%22Avg_Mdcr_Pymt_Amt%22%5D%7D
