## Supplementary material for "Projected healthcare cost savings with transnasal vs. transoral upper gastrointestinal endoscopy in the United States": Non-Human Participant Research

**IRB Administration Office**
87 E. Canfield, Second Floor
Detroit, MI 48201
www.research.wayne.edu/irb

### Notice of IRB Administrative Determination: Non-Human Participant Research (HPR)

**To:** Deepak Gupta, M.D.

**From:** Heather Park-May

**Date:** July 21, 2023

**Re:** Determining National Annual Incidence And Corresponding Costs Of Esophagoscopy Performed Trans-nasally As Compared To Esophagoscopy Performed Trans-orally Per CMS Data 2018-2021

**WSU IRB HPR Number:** **2023 128**

Materials in the above referenced proposal were received by the Wayne State University Institutional Review Board Administration Office via Human Participant Research (HPR) Determination Tool on 07/19/23

This project does not constitute human participant research according to the definition codified in the Common Rule at 45 CFR 46 and FDA regulations. This means that IRB review and oversight is not required for this project. We recommend you keep this memo for your records.

The project also does not involve individuals who would receive a test article (drug or device) as participants and therefore the FDA regulations do not apply. Thus, this project does not require review or approval by the Wayne State University Institutional Review Board.

**Please be aware that projects involving identifiable Personal Health Information (PHI) which the IRB has determined does not meet the definition of human participant research must still adhere to HIPAA regulations, and may require authorization from the covered entity's privacy officer*.**

**Projects must also obtain support from any unaffiliated institution(s) involved.**

**Please contact applicable affiliates: (DMC, Karmanos Cancer Institute, J.D. Dingell Veterans Administration Veterans Medical Center) to obtain authorization to conduct your project.**

Please note that changes to the study plan may impact this determination as to whether the project constitutes Human Participant Research. Please contact the IRB Administration office if there are changes to the study plan that may affect this determination.

Heather Park-May Digitally signed by Heather Park-May
Date: 2023.07.21 13:39:36 -04'00'

Wayne State University IRB Administration Office

### Human Participant Research Determination Tool

According to federal regulations from the Department of Health and Human Services (DHHS) and the Food and Drug Administration (FDA), all human subject/participant research must be reviewed and approved by the Institutional Review Board (IRB). This determination tool is designed to determine whether an activity is considered human participant research under either regulation. The IRB Administration Office can provide assistance with making this determination.

#### Human Participant Research (HPR) Guidance:

Before completing this determination tool, we recommend reviewing the guidance available to help understand how the IRB applies the regulatory definitions to the HPR Determination reviews:

- Does My Study Need IRB Review: Part 1- On- Demand video
- Does My Study Need IRB Review: Part 2- On- Demand video
- HPR Guidance Document

**If you determine that the project does not require IRB review:**

If by the use of this tool, you have determined that the project does not require IRB review, you do not need to submit this form to the IRB office unless you need documentation from the IRB to concur with your self-determination. Add the project title and name of the person completing the project, their title and the date the tool was completed to the first page, and retain this tool in your files to document this determination.

If there are any modifications to your project that could change this determination, please complete this tool again. Submit the appropriate application to the IRB if changes to your project result in the determination that human participant research is involved according to this tool, which is based on the federal regulations.

**If you determine that IRB review is required:**

If you have determined that IRB review is required, IRB approval must be obtained **before** conducting any human participant research activities. Visit the WSU IRB website for additional information and the forms required for a new submission: www.research.wayne.edu/irb . with any questions that come up along the way.

**If assistance is needed, or if written documentation from the IRB office is required:**

Complete the **entire** form and email it to. Include any relevant supporting documents (e.g., grant, protocol, data collection tools). Please do not submit handwritten documents to the IRB office.

**IRB Administration Office**
87 E. Canfield, Second Floor
Detroit, MI 48201
www.research.wayne.edu/irb

**IRB Determination Number (IRB Use Only)**

**2023 128**

#### Section A: Project Staff and Location Information:

Complete all sections of this form if you will be requesting assistance from the IRB Administration Office in making your Human Participation Research determination. Otherwise continue on to Section D to begin the determination tool.

<table>
<tr><td>Project Title:</td><td colspan="4">Determining National Annual Incidence And Corresponding Costs Of Esophagoscopy Performed Trans-nasally As Compared To Esophagoscopy</td></tr>
<tr><td>Name of person conducting the project:</td><td>Deepak Gupta</td><td>Title:</td><td>MD</td><td>Date: 7/11/2023</td></tr>
<tr><td>Status:<br>**Select all that apply**</td><td colspan="4">☑ Wayne State Faculty<br>☐ WSU Graduate Student<br>☐ WSU Undergraduate Student<br>☐ Resident/Fellow/Trainee<br>☐ Other:<br>☑ DMC Staff<br>☑ Karmanos Staff<br>☐ J. D. Dingell VAMC Staff</td></tr>
<tr><td>Division or College:</td><td>Harper-Hutzel Hospital</td><td>Department:</td><td colspan="2">Anesthesiology</td></tr>
<tr><td>Campus Address:</td><td colspan="4">Box No 162, 3990 John R, Detroit, MI 48201</td></tr>
<tr><td rowspan="2">Faculty Sponsor/ Supervisor for this Project:</td><td>Name:</td><td>Title:</td><td colspan="2"></td></tr>
<tr><td>☐ I do not have a Faculty Sponsor/Supervisor</td><td>Phone:</td><td colspan="2"></td></tr>
<tr><td>**Check ALL that apply:**</td><td colspan="4">☑ Behavioral, social, education, non-medical study<br>☑ Medical study</td></tr>
</table>

#### Useful Tips:

1. Carefully read through the definitions provided within the determination tool prior to answering the questions. This will help to ensure an accurate determination is made.
2. If your research involves the use of de-identified data or bio-specimens, include a letter of support from the institution/department providing the data/bio-specimens that confirms that the data/bio-specimens provided will be stripped of all identifiable information before you receive it.
3. If your project requires you to **access**, or **use** identifiable data to complete your data collection, you should check yes to #2.2 in Section D even if the data you are retaining/collecting for analysis will be de-identified.
   a. For example, accessing a patient's medical record to collect data for a chart review gives you access to individually identifiable data even though you have no plans to retain/collect any identifiable data. This is considered a human participant as described by #2.2 under section D of this tool.
4. Review the guidance available on the cover page

#### Section B: Location and Study Data Details:

| | | | |
|---|---|---|---|
| **1.** | Describe the location(s) where activities will take place:<br>CMS 2018-2021 Public Use Files available at https://data.cms.gov/provider-summary-by-type-of-service/medicare-physician-other-practitioners/medicare-physician-other-practitioners-by-provider-and-service/data/ | | |
| **2.** | Select the type of data being collected:<br><br>Check all that apply: | ☑ Secondary or retrospective data collection<br>*(Existing data that was originally generated for reasons unrelated to this project)*<br>☐ Identifiable Protected Health Information (PHI)<br>☐ Prospective Data<br>☐ Secondary or retrospective collection of bio-specimens<br>☐ Prospective collection of bio-specimens | |
| **3.** | Will you require **access** to, or the **use** of any identifiable information pertaining to participants during the process of collecting data? | ☐ Yes<br>☑ No | |
| **4.** | Will data being obtained include individually identifiable information? | ☐ Yes<br>☑ No | If yes, list all identifiers being collected (e.g. name, date of birth, medical record number, email address, other codes; etc.): |

| | | |
|---|---|---|
| **5.** | Could the identities of participants be known to, or be readily ascertained by the investigators? | ☐ Yes<br>☑ No |
| **6.** | Select the source of data collection:<br><br>Check all that apply | ☐ Medical Record Review: Complete #6a.<br>☐ Survey<br>☐ Interview<br>☐ Bio-bank<br>☑ Data Repository<br>☐ Other<br>Describe:<br>CMS 2018-2021 Public Use Files available at https://data.cms.gov/provider-summary-by-type-of-service/medicare-physician-other-practitioners/medicare-physician-other-practitioners-by-provider-and-service/data/ |
| **6a**. | Indicate the institution(s) (Covered Entity) you will be reviewing and collecting medical record data from:<br><br>Check all that apply: | ☐ Detroit Medical Center Facility<br>☐ Karmanos Cancer Institute<br>☐ J.D. Dingell Veterans Administration Medical Center<br>☐ Other<br>Describe:<br><br>☑ N/A- Project does not involve the review of or collection of data from a medical record |
| | **Note:** All applicable institutional policies must be followed at all times. When collecting medical record data outside of your normal responsibilities within the institution, you must obtain approval to access and collect medical record data. | |

| | |
|---|---|
| **7.** | Describe how the data will be obtained (e.g. survey, interview, observation, testing, review of existing records, etc.):<br>CMS 2018-2021 Public Use Files<br>https://data.cms.gov/provider-summary-by-type-of-service/medicare-physician-other-practitioners/medicare-physician-other-practitioners-by-provider-and-service/data/ will be accessed after managing columns and applying advanced filters to export generation of cumulative excel sheets of de-identified providers' data per https://data.cms.gov/resources/medicare-physician-other-practitioners-by-provider-and-service-data-dictionary to compare the following cumulative de-identified providers' data nationally for trans-nasally performed esophagoscopy without anesthesia with or without biopsy as compared to trans-orally performed esophagoscopy with gastro-duodenoscopy potentially with anesthesia with or without biopsy:<br><br>Total Number of Medicare Beneficiaries Nationally<br>Total Number of Services Nationally<br>Total Submitted Charge Amount Nationally By Cumulating Average Submitted Charge Amount Per De-Identified Provider Multiplied With Number of Services Per De-Identified Provider<br>Total Medicare Allowed Amount Nationally By Cumulating Average Medicare Allowed Amount Per De-Identified Provider Multiplied With Number of Services Per De-Identified Provider<br>Total Medicare Payment Amount Nationally By Cumulating Average Medicare Payment Amount Per De-Identified Provider Multiplied With Number of Services Per De-Identified Provider<br>Total Medicare Standardized Payment Amount Nationally By Cumulating Average Medicare Standardized Payment Amount Per De-Identified Provider Multiplied With Number of Services Per De-Identified Provider |

#### Section C: Description of the Project:

**Provide a description of the project with enough detail for the determination.**

| | |
|---|---|
| **8.** | Describe the purpose of this project:<br>The primary purpose of this project is to quantify nationwide total CMS submitted charge amounts and nationwide total CMS allowed amounts for esophagoscopy without anesthesia performed trans-nasally with or without biopsy as compared to nationwide total CMS submitted charge amounts and nationwide total CMS allowed amounts for esophagoscopy (with gastro-duodenoscopy potentially with anesthesia) performed trans-orally with or without biopsy per CMS data 2018-2021 |

| | |
|---|---|
| **9.** | Describe the objectives or aims for the project:<br>The primary purpose of this project is to quantify nationwide total CMS submitted charge amounts and nationwide total CMS allowed amounts for esophagoscopy without anesthesia performed trans-nasally with or without biopsy as compared to nationwide total CMS submitted charge amounts and nationwide total CMS allowed amounts for esophagoscopy (with gastro-duodenoscopy potentially with anesthesia) performed trans-orally with or without biopsy per CMS data 2018-2021 |
| **10.** | Describe how the results will be used/applied:<br>Primarily, total CMS submitted charge amounts nationally and total CMS allowed amounts nationally for esophagoscopy without anesthesia performed trans-nasally with or without biopsy will be averaged out over total number of services performed nationally.<br><br>Concurrently, total CMS submitted charge amounts nationally and total CMS allowed amounts nationally for esophagoscopy (with gastro-duodenoscopy potentially with anesthesia) performed trans-orally with or without biopsy will be averaged out over total number of services performed nationally.<br><br>These totals as well as these averages will be compared for the differences and maybe published as a research report with literature-based exploration of potential reasons for those differences. |

| | |
|---|---|
| **11.** | Describe the participants for the project:<br>CMS 2018-2021 Public Use Files available at https://data.cms.gov/provider-summary-by-type-of-service/medicare-physician-other-practitioners/medicare-physician-other-practitioners-by-provider-and-service/data/ |

#### Section D: Determination Tool:

In order for a project to meet the regulatory definition of human participant (subject) research under HHS regulation 45 CFR 46.102 and FDA regulation 21 CFR 50.3, the project must meet the definition of a human participant(subject) and the definition of research. The following questions will help to determine if your project meets these definitions and requires IRB review.

**Step 1: Does the Project Meet the Regulatory Definition of Research Under HHS Regulations (45 CFR 46)?**

***Research*** means a systematic investigation, including research development, testing, and evaluation, designed to develop or contribute to generalizable knowledge.

**Select all that apply:**

| | | | | |
|---|---|---|---|---|
| 1 | 1.1 | ☐ | Information is expected to expand the knowledge of a scientific discipline or other scholarly field or study and yield one or both of the following: | |
| | | | ☐ | Results are intended to be applicable to a larger population beyond the site of data collection or the specific participants/subjects studied. |
| | | | ☐ | Results are intended to be used to develop, test, or support theories, principles, and statements of relationships or to inform policy beyond the study. |
| | 1.2 | ☐ | The information is collected to share with others in a discipline and is created to make a broad statement (conclusion) about a group of people, procedures, programs, etc. | |

**Step 2: Does the Project Involve Human Participants/Subjects Under HHS Regulations (45 CFR 46)?**

***Human subject*** means a living individual about whom an investigator (whether professional or student) conducting research:

- Obtains information or bio-specimens through intervention or interaction with the individual, and uses, studies, or analyzes the information or bio-specimens; or
- Obtains, uses, studies, analyzes, or generates identifiable private information or identifiable bio-specimens

| | | | |
|---|---|---|---|
| 2. | 2.1 | Does the research involve collecting data through *intervention or **interaction with the individuals? | ☐ Yes:<br>☑ No: |
| | | **Definitions** (45 CFR 46.102):<br>***Intervention includes:***<br>• Both physical procedures by which information or bio-specimens are gathered and manipulations of the participant or the participant's environment that are performed for research purposes.<br>*****Interactions involve:***<br>• Communication or interpersonal contact between the investigator and the participant. Interactions can be in-person or virtual. Examples include interactions via email or social media, or by completing a survey, questionnaire, interview or participation in a focus group. | |
| | 2.2 | Will you obtain, use, study, analyze, or generate *identifiable private information or **identifiable bio-specimens from individuals? | ☐ Yes:<br>☑ No: |
| | | **Definitions** (45 CFR 46.102):<br>***Identifiable private information:** information for which the identity of the participant may be readily ascertained by the investigator or associated with the information.<br>**Private information includes:** | |

| | | |
|---|---|---|
| | | • Information which has been provided for specific purposes by an individual and which the individual can reasonably expect will not be made public (e.g., a medical record, emails, certain listserv communications, class papers and exams, etc.)<br>• Identifiable private information is private information for which the identity of the subject is or may readily be ascertained by the investigator or associated with the information.<br><br>****Identifiable bio-specimen** is a bio-specimen for which the identity of the subject is or may be readily ascertained by the investigator or associated with the bio-specimen. |

| **Step 1 & Step 2 Human Participant Research Determination Decision Tool:** | |
|---|---|
| **Step 1: Research Determination:**<br>• If you checked any boxes in question 1.1 or 1.2, the project is considered research per HHS (45 CFR 46.102(I)) | |
| **Step 2: Human Participant Determination:**<br>• If you answered yes to question 2.1 or 2.2, the project involves human participants per HHS (45 CFR 46.102 (E)) | |
| **Human Participant Research Determination:** | |
| The project is HPR and IRB Review is required if: | Step 1 finds that your project involves research **and** step 2 finds that your project involves human participants.<br><br>• **Submission instructions and guidance are available on the IRB website:** www.research.wayne.edu/irb.<br>• Email your questions to |
| The project is not HPR and does not require IRB review if either of the following applies: | • If step 1 finds that your project does not involve research and/or<br>• If step 2 finds that your project does not involve human participants |
| **Step 3:** Proceed to Step 3 to determine if the project is subject to Federal Drug Administration (FDA) regulations. | |

| **Step 3: Does the activity require IRB review under the FDA Regulations (21 CFR 50)?**<br><br>**Select all that apply** | | | |
|---|---|---|---|
| 3. | 3.1 | ☐ | Activity is conducted in the United States and involves the **use of a drug** in one or more *human subjects (as recipients of a **test article or as controls, patient or healthy 21 CFR 50.3) but is **not** the use of an approved drug in the course of medical practice |
| | 3.2 | ☐ | Activity is conducted in the United States and evaluates the safety or effectiveness of a **device** in one or more *human subjects. |
| | 3.3 | ☐ | Data regarding *human subjects (including controls) will be submitted to or held for inspection by FDA as part of an application for a research or marketing permit. |

| | | | |
|---|---|---|---|
| | 3.4 | ☐ | Data regarding the use of a device on human specimens (including de-identified/anonymous) specimens will be submitted or held for inspection by the FDA as part of an application for a research or marketing permit |
| | **Guidance- Definition (21 CFR 50.3)**<br><br>***Human Subject:** an individual who is or becomes a participant in research, either as a recipient of the test article or as a control. A subject may be either a healthy human or a patient.<br><br>****Test article:** any drug (including a biological product for human use), medical device for human use, human food additive, color additive, electronic product, or any other article subject to regulation under the Food, Drug, and Cosmetic Act. | | |
| | **FDA Human Subject Research Determination Instructions:**<br><br>If any of the boxes in #3 are checked, the activity is human research per FDA regulations and subject to IRB review.<br><br>**Note:** If the activity is determined to be human research subject to FDA regulation, this does not necessarily mean it is also subject to HHS regulation, and vice versa. Separate determinations should be made. | | |

**If assistance is needed, or if written documentation from the IRB office is required:**

Complete the **entire** form and email it to. Include any relevant supporting documents (e.g., grant, protocol, data collection tools). Please do not submit handwritten documents to the IRB office.

#### WSU IRB Determination:

**(To be completed by IRB Administration)**

| ✔ | **Not Human Participant Research - IRB review is not required** | |
|---|---|---|
| | | **Type of activity that does not require IRB review:** |
| | ☐ | Case Report<br>**Note:** IRB approval is required if the case report involves more than three cases |
| | ☐ | Course Related Activities<br>**Note:** IRB Approval is required if a student is involved in an activity designed to teach research methodologies and the instructor or student wishes to conduct further investigation and analyses in order to contribute to scholarly knowledge. |
| | ☐ | Decedents: Research limited to death records, autopsy materials or cadaver specimens.<br>**Note:** IRB approval is required if decedent information contains psychotherapy notes, or information related to HIV, mental health, genetic testing or drug or alcohol abuse |
| | ☐ | Journalism/Documentary Activities<br>**Note:** IRB approval may be required when journalists conduct activities normally considered scientific research intended to develop generalizable knowledge (e.g. systematic research, surveys, and/or interviews that are intended to test theories or develop models). |
| | ☐ | Oral History<br>**Note:** IRB approval is required when the activities are intended to develop generalizable conclusions (e.g., that serve as data collection intended to test economic, sociological, or anthropological models/theories) |
| | ☐ | Program Evaluation/Quality Improvement/Quality Assurance Activities<br>**Note:** Investigators conducting QI/QA projects should ensure that they have received approval from any applicable committees within their department or the site in which the activity will occur |
| | ✔ | Public Use Datasets<br>**Note:** IRB approval is required for the use of restricted use data, if a proposal is required to obtain the dataset, or if a data use agreement is involved. |
| | ✔ | De-Identified Private Information and/or Human Biological Specimens<br>**Note:** IRB approval is required if the information being collected could enable the investigator to identify or readily ascertain the identity of the individual whom the private information or specimens belongs to. |
| | ☐ | Public Health Surveillance<br>**Note:** Activities are limited to those necessary to allow a public health authority to identify, monitor, assess, or investigate conditions of public health importance |
| | ☐ | Other Project type not considered to be HPR<br>Describe: |

| ☐ | **Project is Human Participant Research- IRB Review Required.** | |
|---|---|---|
| | **Type of IRB required:** | |
| | ☐ | Exempt IRB review is required (minimal risk research):<br>Rationale: |
| | ☐ | Expedited IRB review is required (minimal risk research):<br>Rationale: |
| | ☐ | Full Board IRB review is required (greater than minimal risk research):<br>Rationale: |

| | |
|---|---|
| **Reviewer Comments:** | Research: Knowledge gained from this study is not intended to be used to test a theory to gain knowledge about a population. The project is aimed at gaining knowledge about a process,/product/program rather than a human population.(45 CFR 46.102(i)).<br><br>Human Subjects/participants: This project does not meet the regulatory definition of human subjects/participants (46 CFR 46.102(e)) because investigators will be using publicly available data for cost analysis. |
| **Authorized IRB Reviewer Signature:** | Heather Park-May Digitally signed by Heather Park-May Date: 2023.07.21 13:39:53 -04'00' |

**2023 128**
